# Unveiling Associations Between Retinal Microvascular Architecture and Phenotypes Across Thousands of Healthy Subjects

**DOI:** 10.1101/2024.04.05.24305164

**Authors:** Michal Shapira, Smadar Shilo, Yeela Talmor-Barkan, Yaron Aviv, Yotam Reisner, Anastasia Godneva, Adina Weinberger, Alon Skaat, Anat Loewenstein, Eran Segal, Hagai Rossman

## Abstract

Fundus imaging has emerged as a powerful tool, allowing high-resolution and non-invasive visualization of the fundus microvasculature. Recent advancements in artificial intelligence (AI) have enabled the quantification of fundus microvascular characteristics, paving the way for the development of predictive markers for systemic conditions. In this study, we characterize the fundus microvasculature among 8467 healthy individuals, aged 40-70 years. We employ a fully automated segmentation tool to calculate 12 distinct vessel measures, including tortuosity density, vessel density, average width, fractal dimension, distance tortuosity, and curvature tortuosity for both arteries and veins. We provide reference values for these measures by age and sex, and explore the associations between these fundus vessel measures and various clinical features. Our analysis reveals numerous significant correlations, including connections between fundus arterial measures and cardiovascular measurements, lipid profile, sleep apnea indices, glycemic profile, body composition, and smoking status. This comprehensive analysis of fundus microvasculature anatomy in a large healthy population contributes to the growing field of oculomics and highlights the potential of fundus imaging for earlier detection and monitoring of systemic diseases such as metabolic syndrome, cardiovascular diseases and sleep disorders.

## Introduction

High-resolution, rapid, and non-invasive fundus imaging tools have been one of the key developments over the last decades in ophthalmology. Growing evidence supports the ability of fundus imaging to identify systemic conditions as it allows direct and non-invasive visualization of microvasculature in-vivo. Currently, we can quantify fundus microvasculature characteristics by using artificial intelligence (AI) generated segmentations from raw fundus images. These measures can then be used as predictive markers for systemic conditions - a new field called “oculomics” ^1^.

Numerous studies, including some with large population cohorts such as the UK Biobank ^2^, have investigated the associations between fundus microvascular characteristics and systemic diseases. Most of these studies were focused on cardiovascular diseases ^3^, and showed associations between fundus microvascular characteristics and cardiovascular risk factors. Other studies have utilized deep learning from fundus images, in order to directly predict mortality risk ^4^ and conditions such as hypertension, dyslipidemia, and obesity ^3^. Some associations were also found in previous studies between fundus microvascular width, tortuosity, density or fractal dimension and obesity ^5^, blood pressure ^6^, smoking ^7^, sleep apnea ^8,9^, and blood glucose ^3^.

These studies have demonstrated the potential for fundus microvascular measurements to serve as biomarkers for the prediction and diagnosis of systemic diseases. However, these studies are limited, since they tend to focus only on one or relatively few risk factors or conditions, and are examining only one or few fundus vessel measurements. Moreover, the majority of these are based on a small population size and participants with a variety of clinical diagnoses, which cannot represent a healthy population.

In light of the above limitations, the advantages of using fundus imaging at scale for monitoring and identifying systemic health conditions are clear. Accordingly, our study aimed to characterize fundus images that were collected on thousands of healthy individuals as part of the 10K cohort ^10^ collected in Israel during 2019-2023. We quantify a total of 12 distinct vessels measures, including tortuosity density, vessel density, average width, fractal dimension, distance tortuosity, and curvature tortuosity, calculated separately for arteries and veins, using the automated tool AutoMorph ^11^.

This tool leverages deep learning techniques for the automated segmentation of veins and arteries with high accuracy on a large scale. By enabling standard computer vision algorithms to operate on the segmentation, the pipeline extracts global vascular measurements as those mentioned above and thus represents a comprehensive end-to-end solution. In contrast to prior studies that relied on semi-manual tools like QUARTZ or necessitated manual fine-tuning of deep-learning algorithms, this approach is both fully automated and reliable, as it was validated on several independent datasets ^12^.

In the study, we present the distribution of vessel measures and suggest reference values by age and sex, which can serve as a reference for the healthy population- a digital nomogram^1^. Moreover, we analyze vessel measure associations to a comprehensive set of clinical measurements including cardiovascular measurements, lipid profile, sleep apnea indices obtained through a home sleep device, glycemic profile obtained from continuous glucose monitoring (CGM) devices, body composition, and smoking status.

## Methods

### Data

The data was collected as part of the 10K project, a large perspective, longitudinal study. Central view images of both eyes were taken with the iCare DRSplus confocal fundus imaging system (ICare). Center view 45° fundus images were collected without pupil dilation. Body composition measures were obtained by a DXA (Dual-energy X-ray absorptiometry) scan. Body measurements (body mass index (BMI), fat composition and mass, body circumferences), blood pressure and liver ultrasound measurements were collected on the same day. CGM and sleep data (providing sleep apnea and oxygen desaturation indices over 3 nights) were collected in close proximity to the images. Additional clinical data including previous eye conditions and smoking status were collected by the use of electronic surveys as previously described ^10^, and blood test results were uploaded by the participants..

### Study population

Upon arrival at the research site, all participants provided informed consent in accordance with the guidelines set forth by the Declaration of Helsinki. To protect participant confidentiality, any identifying information was removed prior to the computational analysis. The 10K cohort study received approval from the Institutional Review Board (IRB) of the Weizmann Institute of Science. Between 17/02/2021 and 28/03/2023, a total of 9,450 participants in the 10K study had fundus images available, for extraction of vascular measures .Participants with reported diagnoses of glaucoma, cataract, and retinal detachments or breaks were excluded from the study population, along with those with invalid images. Consequently, 8,467 individuals were included. Table S1 summarizes the baseline characteristics of the individuals that had at least one eligible fundus measure.

### Covariates definitions

#### CGM

CGM derived measures are cumulative glycemic features (calculated with the R package iglu ^13^) that were obtained from a two-week-long CGM. We decided to focus on two measurements: estimated A1C (eA1C), which is a transformation of the mean glucose level, and CV, the coefficient of variation of all glucose values.

#### Sleep monitor and sleep staging

The monitoring of sleep in the 10K study utilizes the FDA-approved WatchPAT-300 from Itamar Medical, which employs five sensors for data collection: a wrist-worn actigraph, a finger-worn pulse oximeter and Peripheral Arterial Tone (PAT) probe, and a chest-worn microphone and accelerometer to measure respiratory effort, snoring, and body position. The PAT probe measures changes in the volume of blood vessels by applying consistent pressure to the finger and capturing changes in displaced volume flow using a photoplethysmogram (PPG) sensor. The PAT signal reflects alterations in the autonomic nervous system (ANS) triggered by respiratory disturbances during sleep. The ANS is responsible for controlling essential functions such as blood vessel size and regulation of blood pressure, lung airflow, and the electrical activity and contractility of the heart.

The WatchPAT’s automatic algorithm analyzes the PAT signal amplitude alongside heart rate and oxygen saturation to detect and classify breathing problems during sleep, providing two indices for sleep apnea diagnosis: the Apnea/Hypopnea Index (AHI) and Respiratory Disturbance Index (RDI). The algorithm uses specific signal patterns to calculate these indices, when minimal oxygen desaturation for AHI and RDI events is 3%. The snoring sensor helps clinicians determine if respiratory events are obstructive, while the body position sensor aids in identifying if sleep apnea has a positional component. Additionally, the pulse oximeter calculates the Oxygen Desaturation Index (ODI) by counting the number of detected desaturation events per hour of sleep (minimal desaturation: 4%). The accuracy of these algorithms was validated using gold standard polysomnography (PSG) sleep monitoring ^14^.

The determination of sleep staging was done by combining data from multiple sensors, while sleep/wake detection relies on actigraph data (Hedner et al. 2004) recorded by the device. The accuracy of these algorithms was validated using the gold standard, manually scored polysomnography (PSG) (Hedner et al. 2011).

#### Liver ultrasound measures

Using the Supersonic Aixplorer Mach 30 ultrasound device, we obtain speed of sound measurement and elasticity measurement: the lower the sound speed value, the higher the fat content ^15^, and hence we use the negative sound speed in the analysis to provide positive correlation between the measure (negative sound speed) and fat content. Liver elasticity (based on 2D-ShearWave Elastography) is measured in kPa from ultrasound and serves as an indicator for fibrosis (high values correspond to stiffness and fibrosis).

### Statistical analysis

We used the AutoMorph package ^11^ to calculate 6 different vessel measures: tortuosity density, vessel density, average width, fractal dimension, distance tortuosity and curvature tortuosity (which was taken as the square root of the provided squared curvature tortuosity measure). These measures were calculated separately for veins and arteries (total of 12, described below). We used the square root of the squared curvature tortuosity instead of the raw value to draw its distribution nearer to the normal distribution. Each measure was calculated for each eye separately, and in our analysis we have taken only the mean value. For participants who had more than one imaging, analysis was performed on the first acquired image.

Correlations between these measures and other clinical features, adjusted for age and sex, were calculated using python pingouin package ^16^, and were corrected for multi-hypothesis using Benjamini-Hochberg FDR correction. The percentiles shown in the age-sex reference plots were calculated with a Lowess smoother, using tsmoothie python package ^17^. We next chose 50 measures that were previously shown to be related to the fundus microvascular structure and are measured as a part of the 10K cohort, including: anthropometrics and blood pressure measurements, glucose indicators, thyroid functions, lipid profile, smoking status, measures related to sleep disorders from sleep monitoring and diagnosis of eyes conditions. Only blood tests that were taken in the range of 11 months before the imaging to 3 months after it were included in the analyses. Smoking status and obstructive sleep apnea data were obtained from questionnaires filled by participants at study initiation. All other measures were measured as part of the 10K cohort, the majority being evaluated on the same day.

## Results

### Participant characteristics and study design

Fundus images of 9,450 participants who participated in10K project were analyzed. Images that were ranked invalid by the automatic processing pipeline were excluded, along with images of individuals who didn’t meet the inclusion criteria (see study population section in Methods), resulting in 8,467 individuals included, 52.8% females and 47.2% males, with an average age of 52.3 years and average BMI of 26.1 kg/m^2^ [Table S1]. Fundus vascular measurements were calculated using the AutoMorph package ^11^. As part of the 10K study, measurements of vital signs, anthropometry, blood tests, body composition (obtained by DXA scan - dual energy X-ray absorptiometry), continuous sleep monitoring for 3 nights, and CGM (continuous glucose monitoring) were also collected from the participants. Out of these clinical measurements, we selected 50 that were divided into 6 categories: (1) body composition, (2) cardiovascular state, (3) blood lipid profile, (4) smoking, (5) sleep, and (6) glucose-related measures.

### Reference values of vessel measures extracted from fundus images

12 different vessel measures were calculated for each of the eligible participants in the study population. The measures are described in detail in [Table 2] and are separately calculated for arteries and veins. They include vessel density, the area of blood vessels as a fraction of the total area imaged; average width; fractal dimension, a measurement of vessel complexity; and three different tortuosity measures, which evaluate the “twistiness” or “curviness” of the blood vessels. Percentiles of each measure were calculated using Lowess regression for the entire study cohort, as well as according to age and sex.

**Table 2.** Vascular fundus measures. Mean values for each of the key fundus measures, for arteries and veins separately as well as for females and males; standard deviations are indicated in brackets.

|  |  |  |  |  |  |  |
| --- | --- | --- | --- | --- | --- | --- |
| Measure,<br>mean (SD) | Male (n=4000) |  | Female (n=4467) |  | Description | units |
|  | Arteries | Veins | Arteries | Vein |  |  |
| **Fractal dimension** | 1.353<br>(0.024) | 1.399<br>(0.019) | 1.348<br>(0.025) | 1.394<br>(0.020) | A measure to evaluate the complexity of the vascular branching within the fundus. To calculate this, the box-counting dimension (Minkowski–Bouligand dimension) approach is used <sup>18</sup>. | Unitless<br>(between 1 and 2) |
| **Vessel density** | 0.0387(0.0045) | 0.0497<br>(0.0043) | 0.0387<br>(0.0047) | 0.0496<br>(0.0045) | The area of blood vessels as a fraction of the total area imaged (each measured as sum of pixels) | Unitless<br>(ratio<1) |
| **Average width** | 1.81e5<br>(1.3e4) | 1.92e5<br>(1.3e4) | 1.86e5<br>(1.3e4) | 1.97e5<br>(1.3e4) | Calculated as the ratio between vessels number of pixels and the total length of the vessels | Length<br>[pixels]<br>Can be converted by 1 pixel = 4.3µm |
| **Distance tortuosity** | 3.17<br>(0.91) | 2.56<br>(0.56) | 3.36<br>(1.04) | 2.60<br>(0.59) | A measure of the "twistiness" or "curviness" of a blood vessel as it runs from one point to another. It is calculated by dividing the actual length of the vessel by the straight-line distance between the two points. The measure is averaged across the image<sup>19</sup>. | Unitless<br>(ratio>1) |
| **Curvature tortuosity (the root of squared curvature tortuosity)** | 3.99<br>(1.34) | 3.16<br>(0.88) | 4.26<br>(1.47) | 3.25<br>(0.92) | The curvature of the vessel is measured at a series of points along its length, and these measurements are squared and summed. The resulting value is then divided by the total length of the vessel <sup>19</sup>. | Unitless |
| **Tortuosity density** | 0.693<br>(0.033) | 0.703<br>(0.025) | 0.701<br>(0.035) | 0.708<br>(0.026) | Evaluates vessel tortuosity by summing local contributions to tortuosity by assessing how much each turn curve is different from a smooth curve <sup>20</sup> | 1/length<br>[1/pixels]<br>1 pixel =<br>4.3µm |

### Relationship between fundus vessels measures

To analyze the association between different extracted measures, we calculated pairwise Pearson correlations and used hierarchical clustering to group them [Figure S1]. A threshold of +-0.7 was used to determine which measures belonged in the same cluster, consistent with the first or second degrees of hierarchy grouping. This resulted in seven clusters among the 12 measures studied: (1) vessel density and fractal dimension, (2) artery average width, (3) vein average width, (4) artery tortuosity density, (5) artery distance tortuosity and curvature tortuosity, (6) vein tortuosity density, and (7) vein distance tortuosity and curvature tortuosity. Measures with similar characteristics tended to cluster together (e.g. vessel density and fractal dimension, tortuosity density, and distance tortuosity). In order to further examine the fundus vascular measures, we selected one representative measure from each cluster, with the exception of artery fractal dimension and artery vessel density, which had a strong correlation (0.88) but are often studied separately in previous research publications. Therefore, both were included in our further analysis. Furthermore, as the three tortuosity measures are intended to assess similar phenomena and have been compared in earlier studies ^20^, we chose to include only tortuosity density measures in our analysis. Still, the other measures were accounted for while adjusting p-values. To conclude, the studied measures for further analysis were: (1) artery vessel density, (2) artery fractal dimension, (3) average artery width, (4) average vein width, (5) artery tortuosity density, and (6) vein tortuosity density. The p-values were calculated for all measures, and they were all taken into account while adjusting for multiple hypotheses.

### Progression of fundus vessels measures with age

In order to evaluate the progression of various measures with age, percentiles were calculated for the entire study population [Figure 2]. We found variability in the way the fundus vessel measures change with age. Our analysis revealed that more than half of the selected measures demonstrated significant correlation with age that exceeded 0.1 in absolute magnitude. Specifically, the fractal dimension of arteries tended to decrease with age (r = −0.17, p<10^−35^), as did the average width of arteries (r = −0.2, p<10^−87^) and the density of arterial vessels (r = −0.25, p<10^−88^). Nevertheless, the manner in which these measures decreased varied; The decrease rate in the average width of arteries moderated with age (there was more decrease per year from age 40 to 50 than from 60 to 70), while the decrease rate in the density of arterial vessels and fractal dimension became more pronounced as the age progressed. Conversely, tortuosity increased with age, as evidenced by the correlation between age and both artery and vein tortuosity density (r = 0.11, p<10^−18^; r = 0.09, p<10^−13^, respectively). Although all the correlation patterns were of similar direction in both females and males, the precise change rate varied between males and females, as is described in [S4].

**Figure 1:**
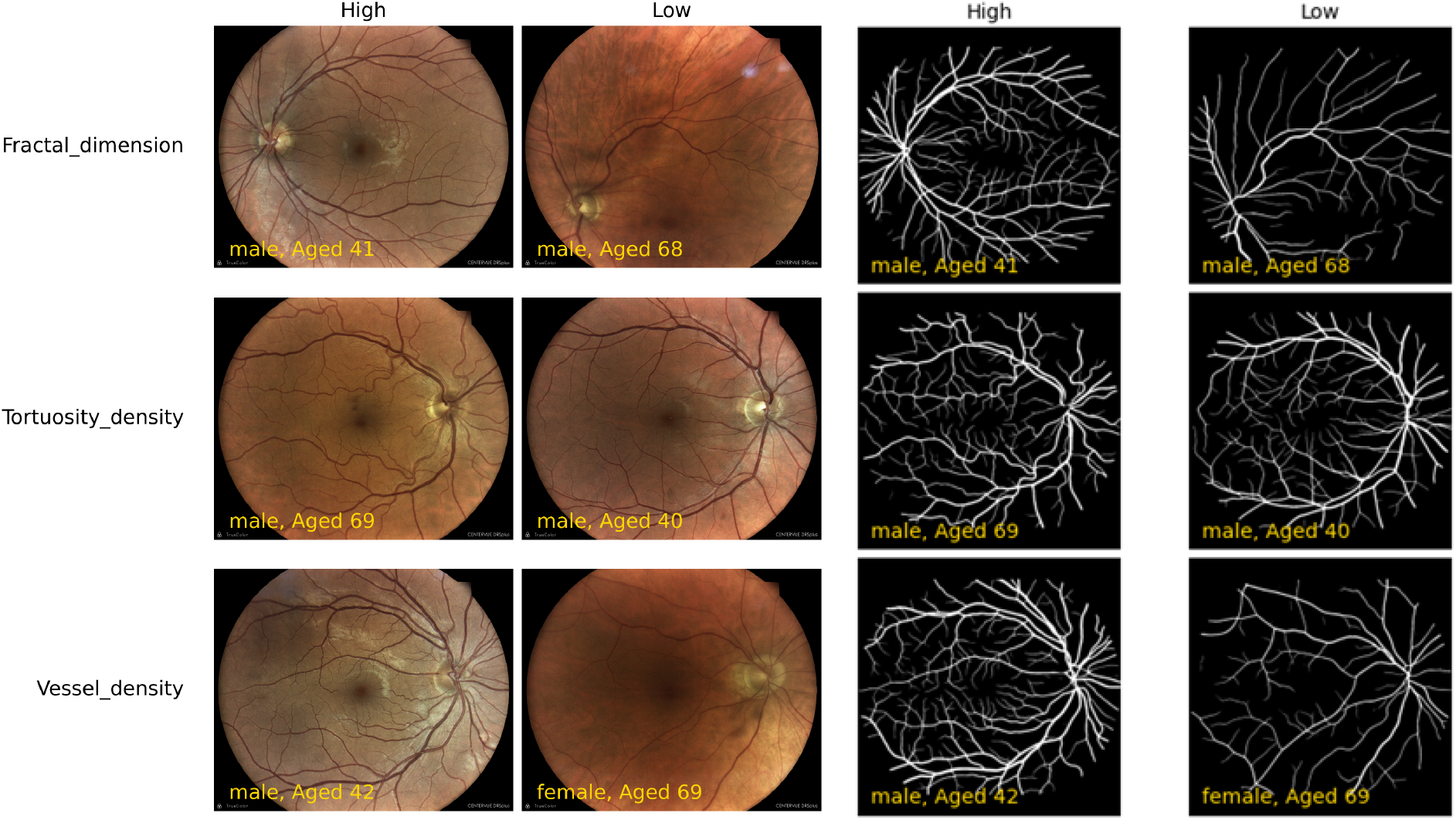
Imaging data segmentation process and key fundus measures across age and sex. Each row presents examples of the named fundus measure with high and low values, in different ages and sexes. **A:** Original images: **B:** Images after segmentation. Fractal dimension and vessel density are typically lower in older age, while tortuosity rises with age.

**Figure 2:**
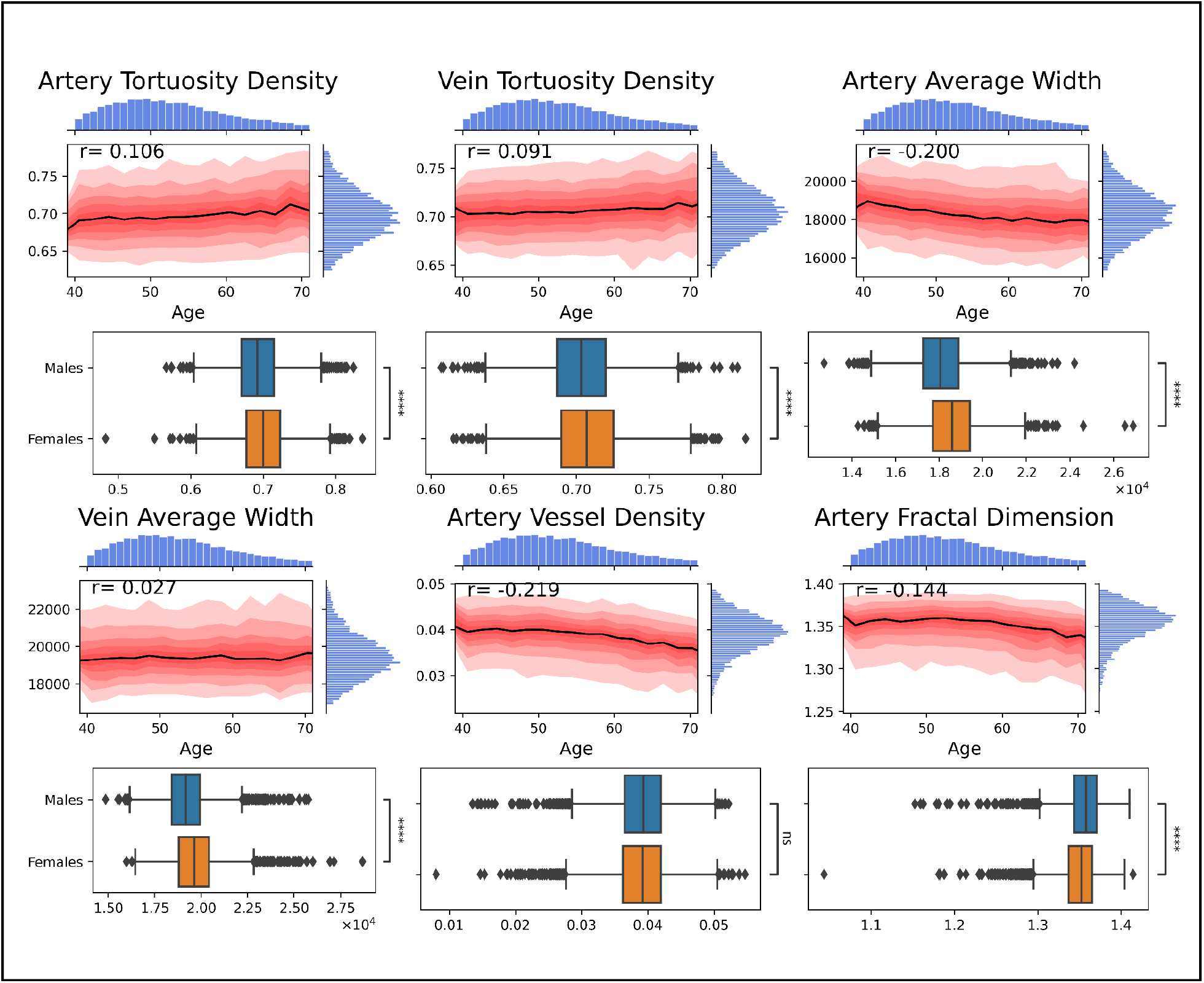
Age and sex reference plots for key fundus measures. The upper plot of each measure presents the median (black line) and percentile (10%-90%, in red) values of the fundus measure titled, by age (x-axis, in years). The bottom plot presents the distribution of the fundus measures (x-axis) by sex. Tortuosity measures increase moderately with age; artery average width, fractal dimension and vessel density decrease with age; vein average width is not correlated with age. Tortuosity and average width are higher among females; artery vessel density is higher among males.

### Association with sex

Artery fractal dimension was significantly higher in males (p<10^−23^)[Figure 2]. Artery (p<10^−80^) and vein (p<10^−53^) average width, and artery (p<10^−22^) and vein (p<10^−11^) tortuosity density were significantly higher in females. Artery vessel density was the only one of the six fundus measures that did not seem to differ by sex.

### Correlation of fundus vessel measures to other clinical measures

The associations between tortuosity density, vessel density, average width, fractal dimension, and squared curvature tortuosity in both veins and arteries with 50 clinical measures from the 10K cohort were analyzed while adjusting for age and sex by addressing them as covariates and calculating partial correlations. The 50 measures were then divided into 6 categories: (1) body composition, (2) cardiovascular, (3) blood lipid profile, (4) smoking, (5) sleep, and (6) glucose related measures. Figure 2 displays the correlation coefficients for representative clinical measures with the 6 key fundus measures defined earlier.

All the clinical measures in the body measurements category have shown a similar correlation pattern with artery vessel density (orange in Fig. 3) and artery average width (green in Fig. 3) - both were decreased in higher BMI, waist to hips ratio, and fat percentage composition. Liver negative sound speed, serving as an indicator for steatosis, was also negatively correlated with artery vessel density and average width. Fractal dimension width (brown in Fig. 3) was similarly significantly negatively correlated to body composition measurements - waist-to-hip ratio and BMI. Vein average width (purple in Fig. 3) showed the opposite correlation patterns, being positively correlated with all the body measurements except for liver elasticity (which was not significantly correlated with any of the fundus measures).

**Figure 3:**
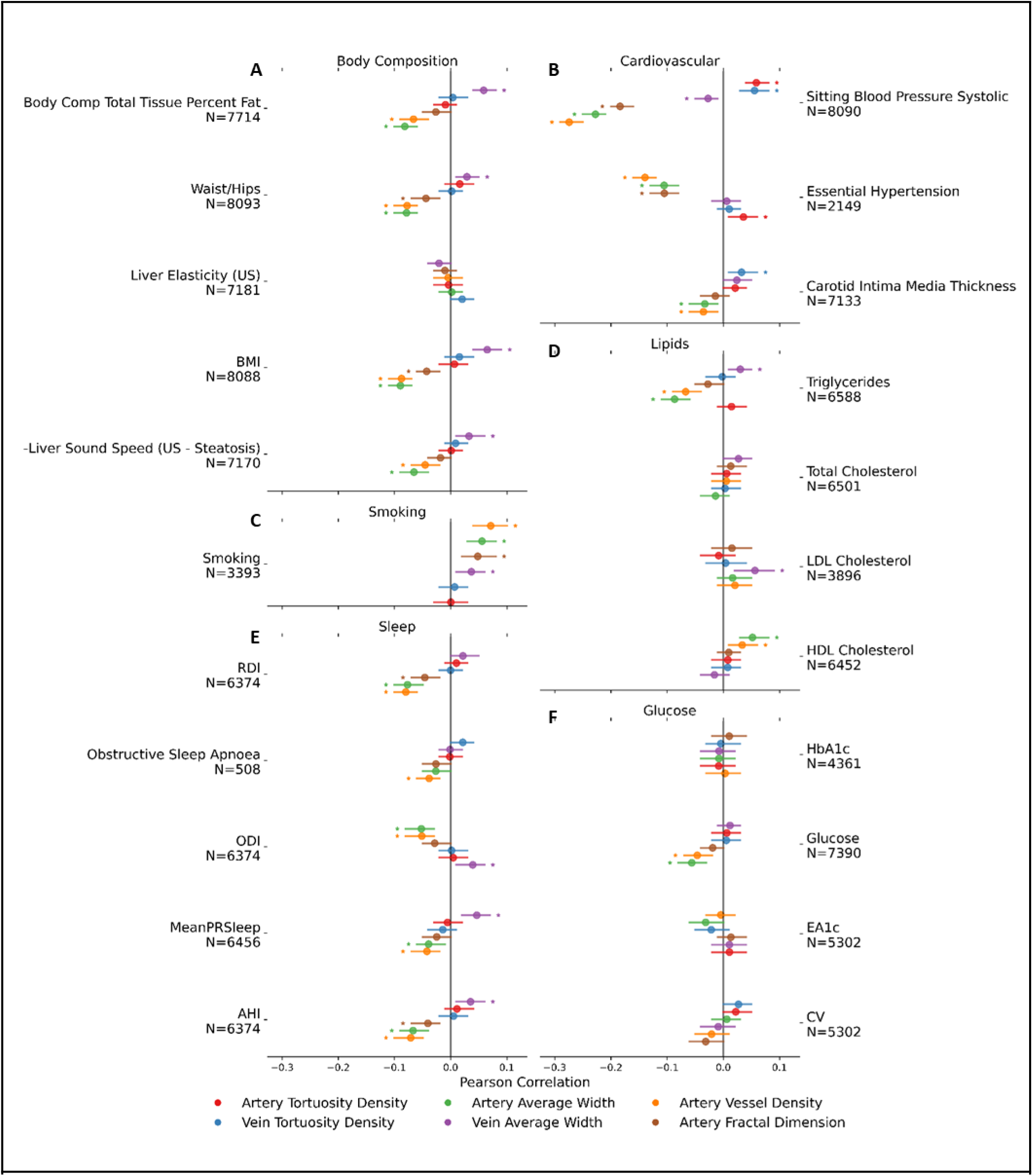
Correlations of fundus vascular measure with clinical measures adjusted forage and sex. Dots represent the correlation coefficient between the clinical measure labeled on the y-axis and the fundus measure of the corresponding color. **A:** Body Composition: artery vessel density, fractal dimension, and average width are negatively correlated with most clinical measures; vein average width is positively correlated. **B:** Cardiovascular: artery vessel density, fractal dimension, and average width are negatively correlated with most clinical measures; both vein and artery tortuosity are positively correlated. **C:** Smoking: artery vessel density, fractal dimension, and average width are positively correlated with smoking. **D:** Lipids: HDL cholesterol is positively correlated with artery vessel density and average width, while triglycerides are negatively correlated with both; LDL cholesterol is positively correlated with vein average width. **E:** Sleep: Vein average width is positively correlated with sleep measure; artery vessel density, fractal dimension, and average width are negatively correlated. **F:** Glucose: Glucose is the only measure of the category that is associated with fundus measure, being negatively correlated with artery fractal dimension and vessel density

In the cardiovascular category, which includes measures of blood pressure (both direct measures and hypertension self-report by questionnaire), and carotid intima-media thickness, all significant correlations were always with the same consistent direction. Artery fractal dimension, artery and vein average width, and artery vessel density were negatively correlated with blood pressure (e.g., low fractal dimension is correlated with high blood pressure). Tortuosity (both artery and vein tortuosity density) was positively correlated with blood pressure. All the mentioned correlations were significant in all direct blood pressure measures correlations. ECG findings such as high voltage in leads I and AVL (mv) as well as P length (ms) were found to be negatively correlated with artery fractal dimension, average width and density, and positively correlated with artery tortuosity density. When excluding patients with high systolic blood pressure (>140) from the analysis, these parameters were no longer correlative to fundus findings. As these ECG findings strongly correlate with hypertension (eg. surrogate markers of atrial size and left ventricular hypertrophy), this further strengthens the correlation between fundus findings and hypertensive target organ damage.

In the lipids category, where all the measures are based on blood tests, a positive correlation was found between HDL cholesterol and artery width and density, with no other fundus association to HDL. LDL cholesterol showed different correlation patterns, positively correlating with only vein average width. When taken together, total cholesterol was not found to be associated with any of the fundus measures. Triglycerides were significantly negatively correlated to arteriolar density and width, while significantly positively correlated to vein average width.

Interestingly, the density of vein and artery tortuosity - which were positively correlated with high blood pressure - were negatively correlated with current smoking. Furthermore, artery average width, fractal dimension and vessel density, which were negatively correlated with blood pressure and metabolic measures like BMI and fat composition - were positively correlated with smoking. Vein average width was also positively correlated with smoking.

In the sleep category, we analyzed correlations with mean sleep pulse rate, ODI (Oxygen Desaturation Index), RDI (Respiratory Disturbance Index), and AHI (apnea-hypopnea index), all obtained from continuous sleep monitoring during a 3-night home sleep apnea test, and self reported obstructive sleep apnea. Artery vessel density and fractal dimension were significantly negatively correlated with all the analyzed measures. Likewise, artery average width was also significantly negatively correlated with all the sleep measures except for the mean pulse rate (ODI, RDI, AHI, obstructive sleep apnea).Vein average width was showed opposite correlation patterns, being positively correlated with ODI, AHI and mean PRS.

In the glucose category, while eA1C was not significantly correlated with any of the fundus measures, four fundus measures were correlated with CV: artery vessel density and fractal dimension were both negatively correlated with the coefficient, while both vein and artery tortuosity density was positively correlated with it.

## Discussion

In this study, we present a comprehensive analysis of a distinctive data set of fundus images, wherein multiple indices characterizing the fundus vasculature were extracted. With over 8400 unique individuals, this work stands as the first large-scale research that incorporates the automatic extraction of 12 distinct vascular measures. As a result, it is poised to serve as a fundamental reference for future research on fundus vascular characteristics, their correlation to different body systems and functions and as a reference map and nomograms for analysis of individuals since the software used is freely available for all^2^.

Analyzing the 12 fundus measures, we have found that some are highly correlated with one another, and could be clustered into 6 groups; accordingly, we decided to focus on 6 representative measurements. By examining the associations between the fundus vascular measures and both age and sex separately, we identified varying patterns in the progression of fundus vascular measures with age. **Artery fractal dimension** decreases with age, as the complexity and branching of the fundus vascular network decrease, in accordance with findings in previous studies ^21–24^). **Artery average width** also decreases with age; factors suggested to contribute to this association include the accumulation of fatty deposits and the development of plaque and a stiffening of the arteries, which can result in reduced vascular width as the vessels lose their ability to expand and contract efficiently ^25^.

On the other hand and in contrast to several previous studies that observed a narrowing ^26^ or widening ^27^ of veins with older age, our findings indicated that the **average width of veins** remained unchanged with age, in line with the work of ^28,29^) and (^28,29^ Both **arteriolar and venular tortuosity density** increased with age, as indicated in previous studies ^27^. **Artery vessel density** decrease with age was the most pronounced, in accordance with previous smaller studies ^23,30,31^.

Moreover, we found different associations with systemic health indicators measurements for each fundus measure. The most noteworthy and interesting findings of the current study include the following: **Artery tortuosity density** increases with blood pressure measures, carotid intima-media thickness, and hypertension, as well as correlated with CV index extracted from CGM. Previous studies have found supporting evidence of the association for blood pressure, although the tortuosity was measured in various ways ^3,32^, and suggested that augmented fundus vascular tortuosity could potentially correlate with heightened blood flow and angiogenesis. A decline in fundus vascular tortuosity was hypothesized to be associated with endothelial dysfunction, thereby impairing perfusion or oxygenation within the microvasculature ^33^. Likewise, **vein tortuosity density** was found to be positively correlated with blood pressure and CV index, and was also found to increase with body measures including waist to hips ratio, body fat percentage, and BMI, and with sleep apnea indices such as RDI, ODI, OSA (Obstructive Sleep Apnea), and AHI. Our results are aligned with previous small-scale studies that demonstrated higher tortuosity in participants with obstructive sleep apnea ^34^. In addition, **artery average width** was negatively correlated with blood pressure (all related measurements) and body measurements including waist to hips ratio, body fat percentage, BMI and liver steatosis. Association between fundus artery atherosclerotic lesions and nonalcoholic fatty liver disease has been previously shown ^35^. Studies have demonstrated that narrower fundus arterioles are strongly correlated with elevated ambient BP, and less strongly with prior BP levels ^36^. Arterial average width was also negatively correlated to glucose, associating the narrowing of arteries with a rise in glucose levels, a finding that is in line with previous findings indicating that fundus arteriolar narrowing predicts incidence of diabetes ^37^. The measure was also negatively correlated with most sleep apnea indices: RDI, OSA, ODI and AHI, in agreement with a previous smaller study that found it correlated to AHI ^38^. Additionally, arterial width was found to correlate with smoking, which will be discussed later in this section.

**Vein average width** was similarly negatively correlated with direct blood pressure measurements but to a much lesser degree than its parallel artery measure. However, its correlation with body measurements showed an opposite direction compared with the artery’s width - it was positively correlated with BMI, waist to hips ratio, and liver steatosis. Interestingly, the vein average width is positively correlated with body measurements but negatively correlated with blood pressure as compared to other fundus vascular measurements, which showed agreement in the correlation direction between these two categories. Similar to the body measures correlations, its correlation with direct measurements of sleep indices - ODI, AHI, and mean pulse rate - were positive. Overall, although vein and artery width have a r=0.67 correlation, they present opposite correlation patterns in some cases: body measures such as BMI and liver fat are negatively correlated with arterial width but positively correlated with venous width; sleep indices such as ODI and AHI are also negatively correlated with arterial width but positively correlated with venular width. Several mechanisms were proposed to explain metabolic conditions’ effect on venous dilation. For example, Dyslipidemia, or abnormal lipid levels in the blood, is a feature of metabolic syndrome and of hyperglycemia, and can contribute to endothelial dysfunction ^39^, which can affect the ability of the veins to dilate properly. Additionally, obesity might predispose individuals to venous stasis, which can contribute to venous dilation ^40^.

The predictive value of arteriolar and venular width on cardiovascular events has been widely reported. Atherosclerosis Risk in Communities (ARIC) study, a US cohort of over 10,000 individuals, was one of the first studies to quantitatively measure arteriolar and venular calibers, and reported that narrower arterioles, represented as lower arteriole to venule ratio (AVR), predicted 3-year risk of coronary heart disease (CHD) events. This association was found only in women, not in men ^41^. The Blue Mountains Eye Study (n = 3654) showed separated prediction of arterial narrowing and venular widening. They showed that wider fundus venules predicted 9-year risk of CHD death in both men and women without a history of pre-existing CHD, while fundus arteriolar narrowing additionally predicted CHD death in women ^42^. A meta-analysis of 21428 individuals showed that narrower fundusl arterioles and wider fundusl venules were associated with an increased risk of CHD in women but not in men ^43^.

**Artery vessel density** was the most correlated (in absolute value, out of all the fundus measures compared) with blood pressure measurements and body measurements, negatively correlated to all of them (except liver elasticity which wasn’t significantly correlated with none of the eye measures). Moreover, it also had one of the two highest correlations with each of the sleep indices - they decrease as the vessel density increases, a pattern that was shown previously in a small study examining obstructive sleep apnea ^34^. Negatively correlated with also CGM’s mean glucose, this measure is placed as one with a potential to indicate many systematic conditions.

**Artery fractal dimension** was found to be highly correlated with artery vessel density, which explains why the two measures share many correlation patterns: sleep indices such as AHI and RDI, triglycerides, smoking status, cardiovascular, and body measures. Nonetheless, arterial vessel density is noticeably more correlated with all indices and is associated with glucose, which was not found to be associated with fractal dimension. Current smoking status was positively correlated with artery fractal dimension, vessel density, and artery and vein average width; was opposite in correlation to the correlation of blood pressure and sleep indices. Nevertheless, these results are consistent with a previous study ^7^ that reported a positive correlation between fractal dimension and the number of pack-years. A possible explanation is a pathological angiogenic effect, mediated by multiple growth factors, including vascular endothelial growth factor. Another explanation that the authors suggested is the existence of a more complex vascular network that may compensate for the decreased blood oxygen-carrying capacity of smokers and decreased blood-O2 affinity in order to meet the fundus high oxygen demand. In any case, these results underscore the importance of further mechanistic investigations into the impact of smoking on microcirculation.

In collective consideration, our findings establish compelling negative associations between arterial width, density, and fractal dimension with diverse metabolic and cardiovascular conditions. These findings highlight the potential of these fundus vessel measures as indicators for further cardiovascular profiling and suggest the promising role of fundus imaging as a potential biomarker for the detection and assessment of metabolic and cardiovascular disorders.

### Limitations of the study

One of the study limitations is the accuracy in calculating the fundus vascular measurements. First, all the quality control processes were conducted automatically, which can lead to several images being inadequate as they are not validated manually. Second, although the AutoMorph pipeline performed well on several validation datasets, we didn’t further test its accuracy on our data. Additional research is needed to gain a better understanding of the biological and mechanistic implications of the associations identified in this study between the fundus vascular measures and clinical indicators

## Data Availability

Data will be available by cloud platform to researchers by request.

## Supplementary Appendix

**Figure S1:**
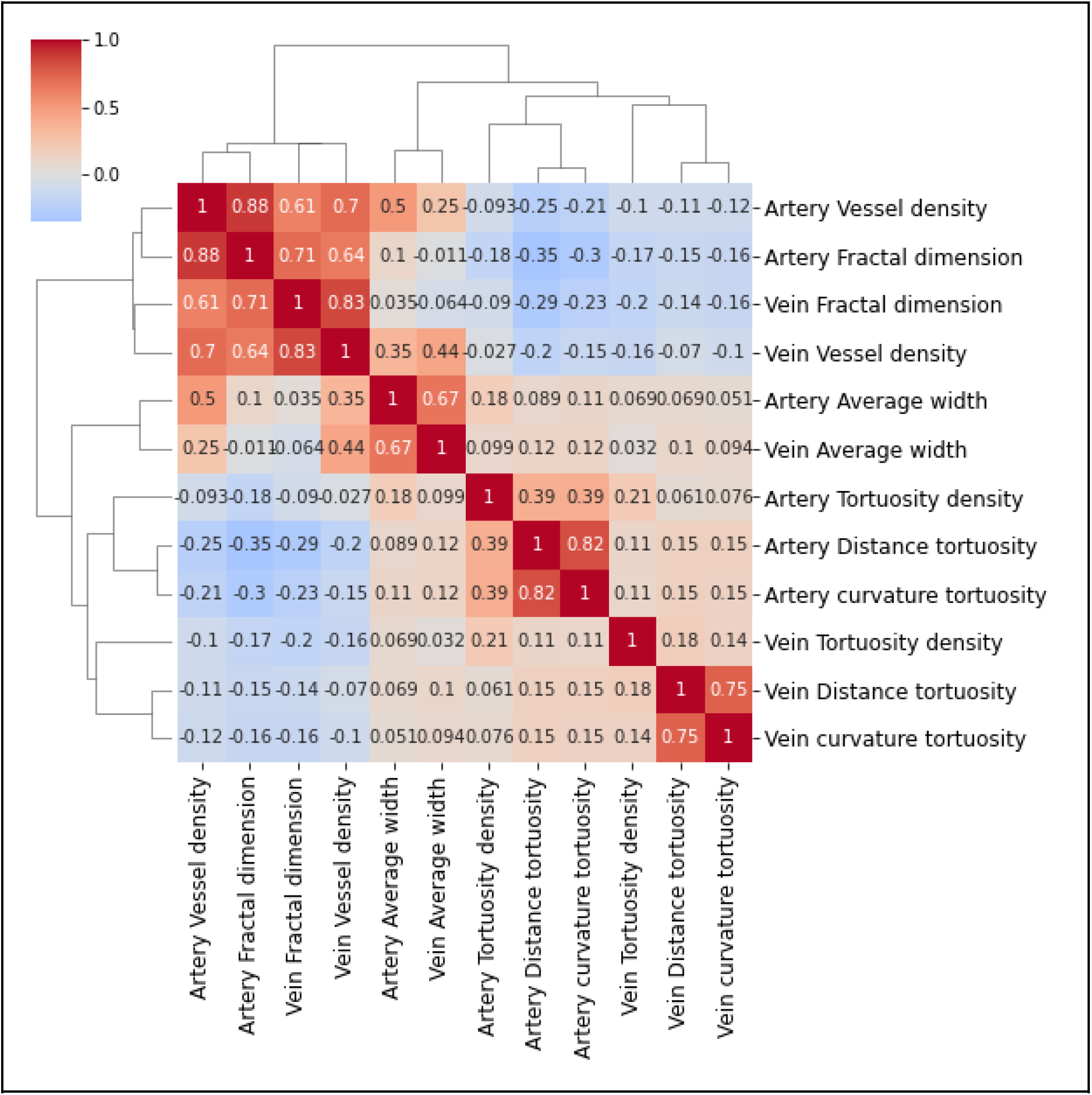
Correlations between all fundus vascular measures, clustered with hierarchical clustering. The numbers and colors represent the correlation coefficient between the pair of fundus measures. A threshold of r=0.7 was set to cluster the measures into 6 clusters.

**Table S1.**
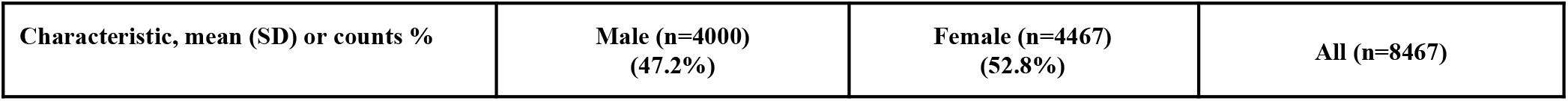
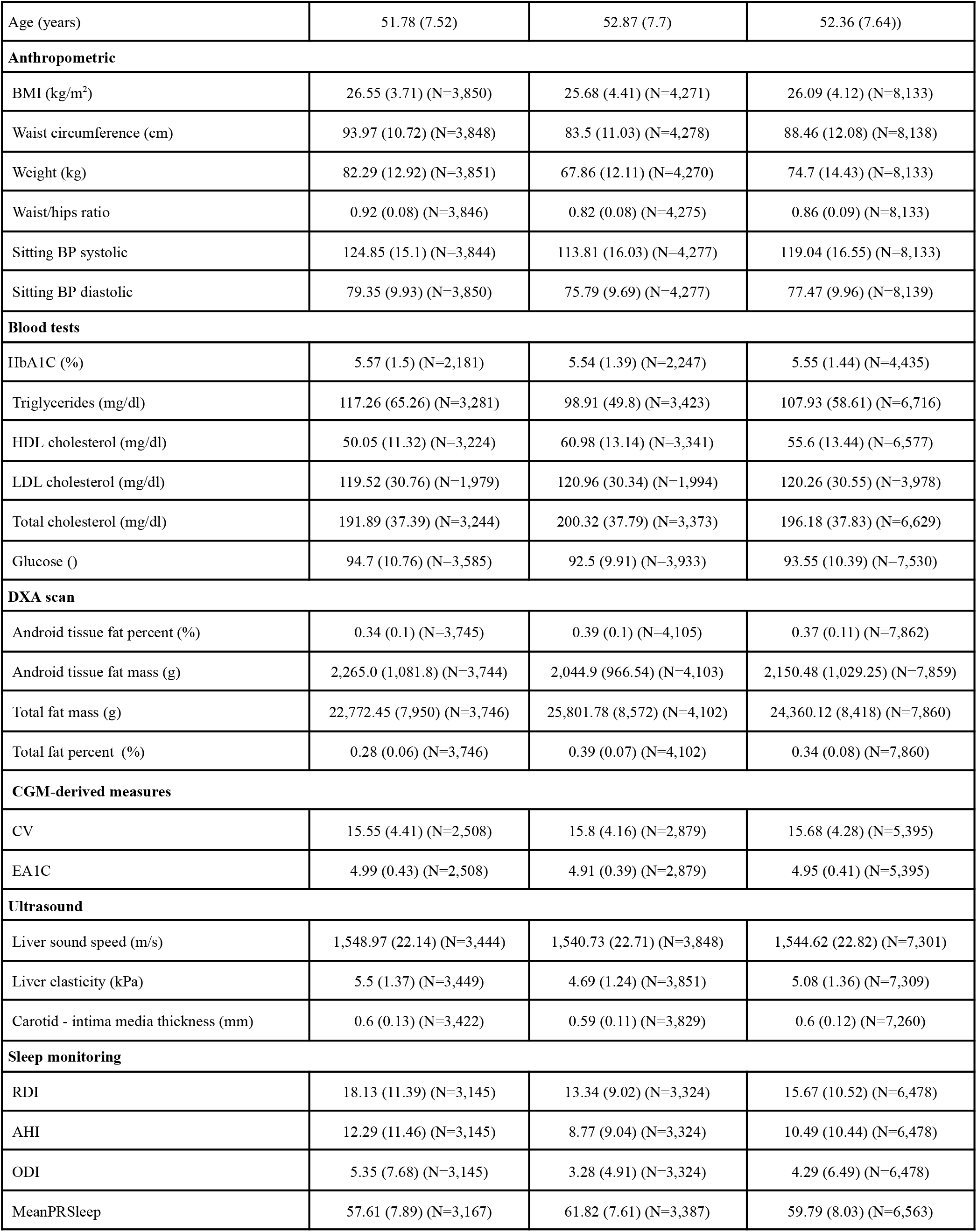
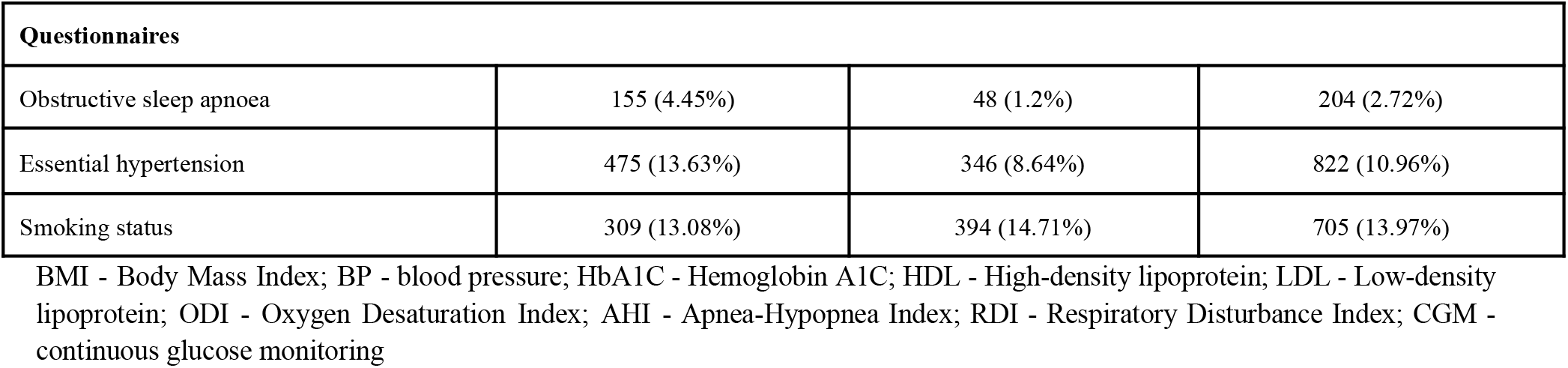
Baseline characteristics of the study population.

**Figure S2:**
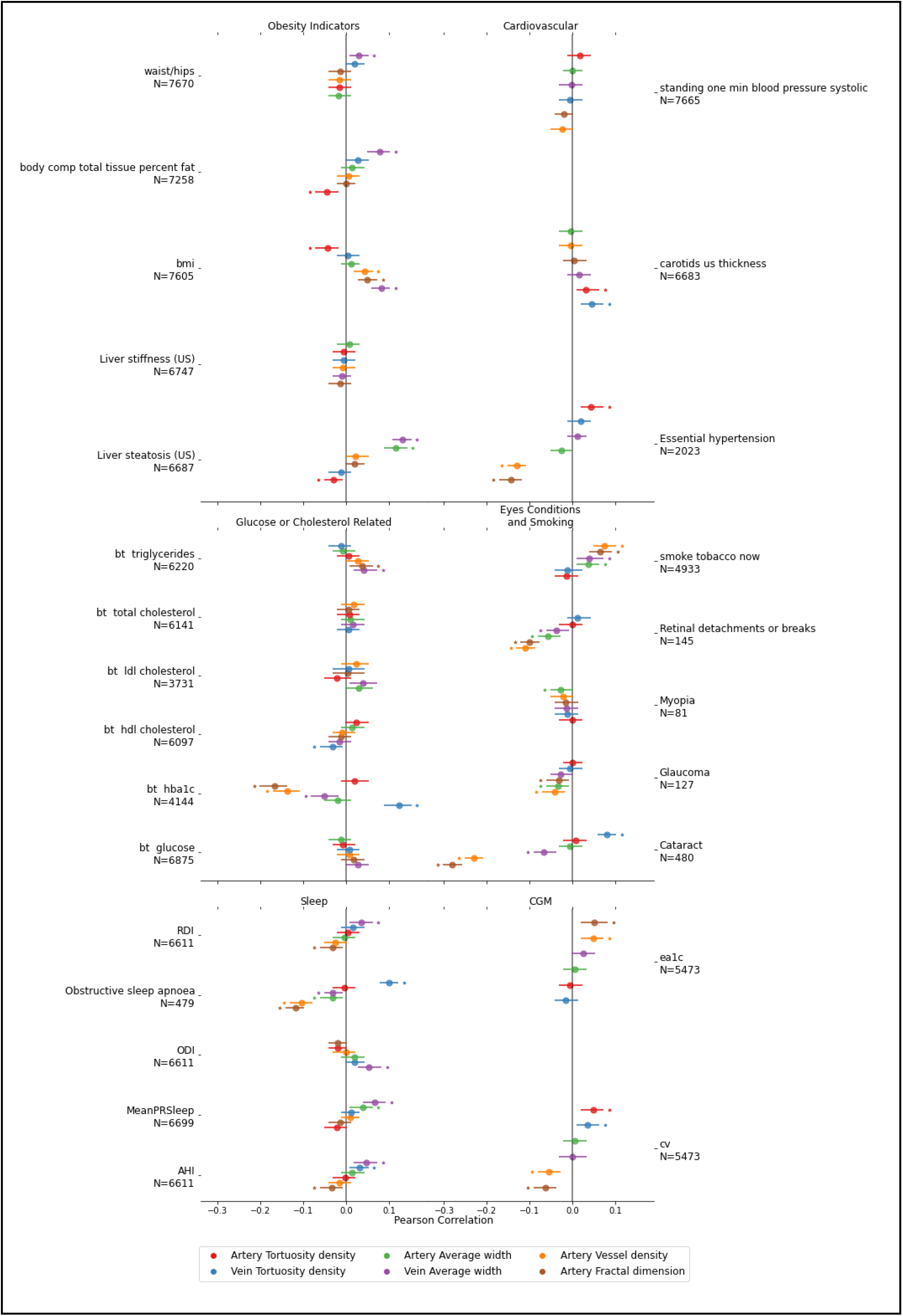
Correlations adjusted for age, sex, and 4 blood pressure measures: sitting and lying blood pressure (both systolic and diastolic).

### Correlation between the left and right eyes

The mean difference between the eyes of an individual is up to 2.5% in all of the extracted measures. In 95% of the participants, the difference between the eyes in the fractal dimension is below 4%, and in average width, vessel density, and tortuosity density below 13%. Distance tortuosity differences are much higher, and 95% are lower than only 89%.

**Figure S3:**
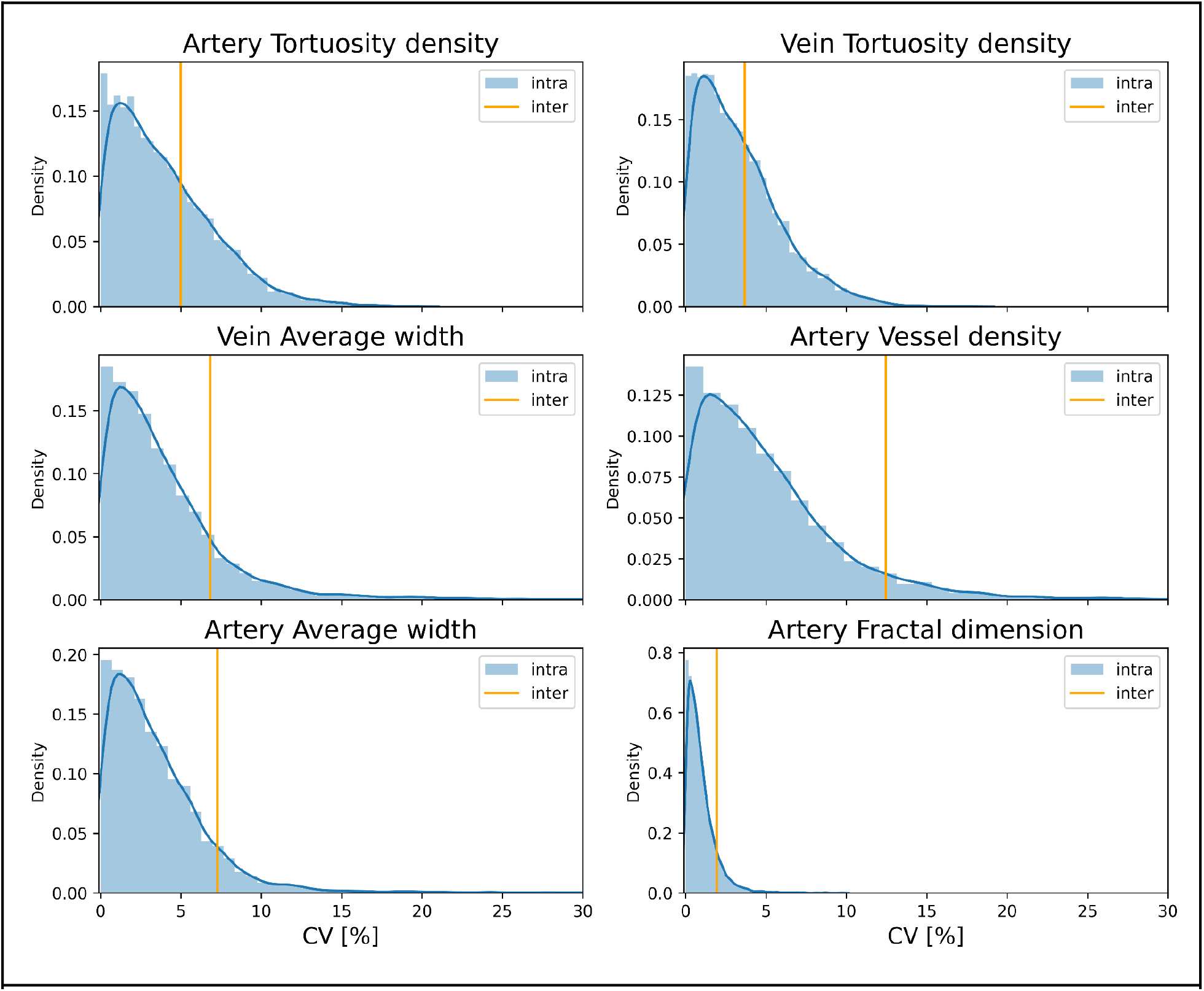
Inter and Intra coefficient of variation in fundus measures. In blue - the coefficient of variance between images of left and right eyes for each individual. In orange - the coefficient of variance of the mean feature value among individuals. In all measures except the tortuosity ones, the intra-variance is significantly lower the inter-variance. In tortuosity measures the typical intra-values are also lower than the inter ones, but to a lesser extent. The lower intra-variance in all the measures enables the use of the mean value of left and right eyes.

**Figure S4:**
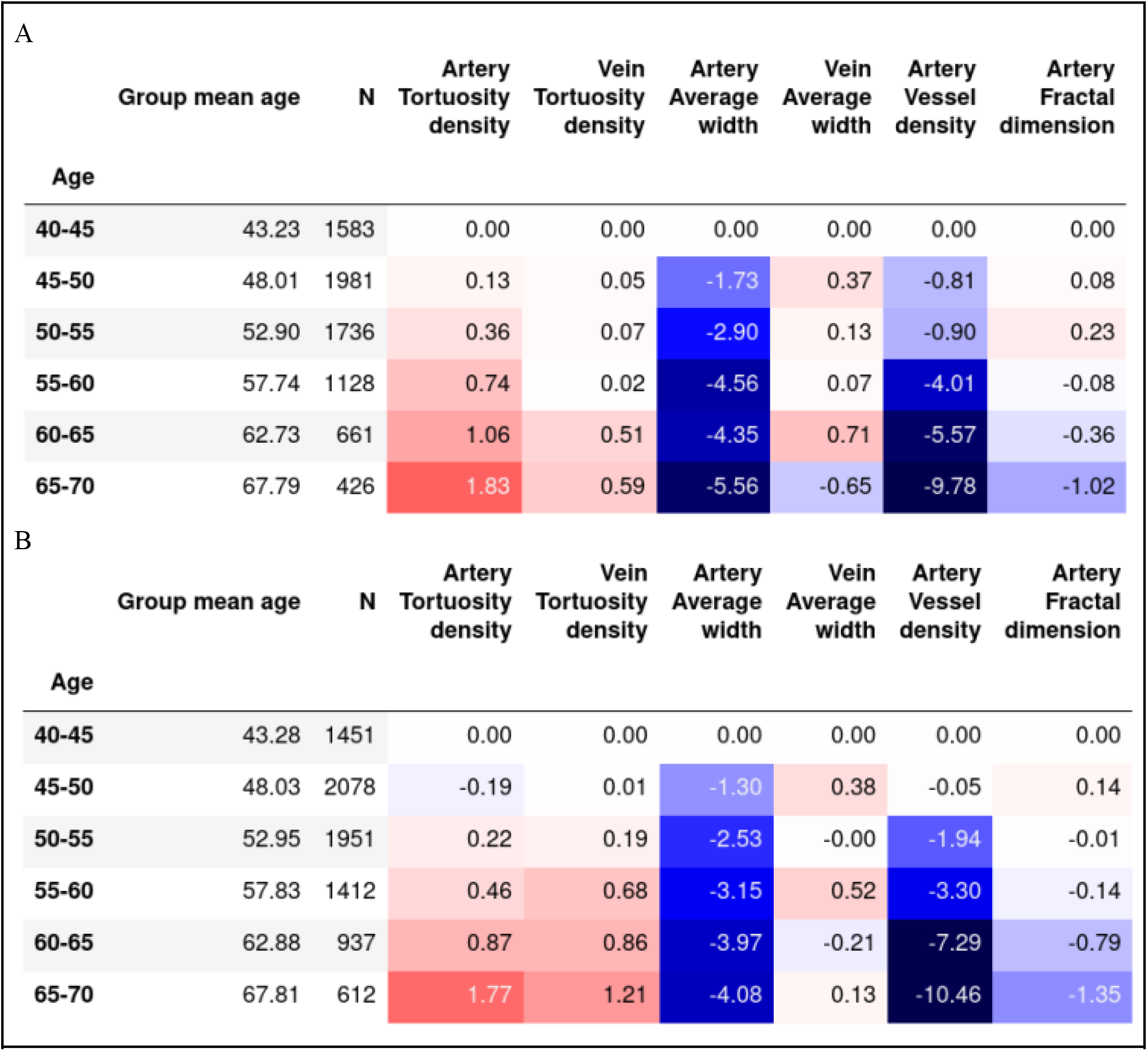
Percentage change in mean values of fundus measures per age relative to mean values at age 40-45 years. **A**: in males. **B:** in females.

## Footnotes

1 Available at https://github.com/michalshap/BeyondFundus

2 Available at https://github.com/michalshap/BeyondFundus

